# Automated Identification of Contextually Relevant Biomedical Entities with Grounded LLMs

**DOI:** 10.1101/2025.07.07.25331004

**Authors:** Manuel Watter, Claudia Giuliani, Gita Benadi, Felix Engel, Harald Binder, Klaus Kaier

**Affiliations:** Institute of Medical Biometry and Statistics, Medical Faculty and Medical Center, University of Freiburg, Freiburg, Germany; Center for Integrative Biological Signaling Studies (CIBSS), University of Freiburg, Freiburg, Germany

## Abstract

This study investigates the effectiveness of different large language models (LLMs) for automated biomedical entity annotation in research articles with a focus on contextualized and grounded results. A 4-step generative workflow iteratively generates and refines entity candidates by considering a metadata schema for context and agentic tool use for validation with the PubTator 3 data base. The precision of this flow was assessed with a random effects meta-analysis after face-to-face interviews with authors of six papers from the Collaborative Research Center (CRC) 1453 “NephGen”. With an overall precision of 91.3%, the selected models provide qualitatively valuable annotations, with models GPT 4.1, GPT-4o Mini, and Gemini 2.0 Flash showing the highest precision. While GPT 4.1 and Gemini 2.0 Flash excelled in the total number of correct annotations, GPT-4o Mini and Gemini 2.0 Flash were fastest and most cost-effective. Large variations in annotation count and the conflation of publication and dataset-specific annotations highlight that human review (“human-in-the-loop”) is still important. The results further highlight the trade-offs between precision, total number of correct annotations, cost, and speed. While quality is paramount in collaborative research settings, cost-effectiveness could be more critical in public implementations.

## Introduction

Long-term research institutions such as Germany’s Collaborative Research Centers (CRCs) rely on structured data sharing and interoperable metadata to ensure long-term data reusability, in line with the FAIR principles [1]. In biomedical research, extracting entities such as organisms, cell lines, or genes from datasets and publications to create searchable tags improves discoverability and reuse. Because manual annotation is time-consuming, automated approaches such as named entity recognition are employed to identify and classify terms (e.g.. tagging “mouse” as an “organism”) [2]. Large Language Models outperform traditional approaches due to their ability to consider context in an unstructured text [3, 4].

Determining “how many” or even “which” entities should be extracted is ultimately an ill-posed question: relevance is shaped by the scientific context and the purpose of the downstream analysis. Moreover, entity relevance shifts with the experimental focus: cell-line names, for instance, are crucial when comparing in-vitro assays, but may be noise for population-level genomic studies. Consequently, optimal entity selection becomes a moving target that must be tuned to the downstream tasks. In our case, we have established a metadata schema in coordination with the experts of and for the CRC [5]. Until now, scientists had to manually annotate their data sets using this schema [6]. The schema will now serve as contextual guidance for the LLM as to,,how many” and,,which” entities to consider.

In recent years, there has been a shift from fine-tuning to in-context learning (ICL) as key strategies for adapting Large Language Models (LLMs) to downstream tasks. ICL guides the model using examples embedded directly in the input prompt, without modifying its internal parameters [7]. In contrast, fine-tuning updates the model’s parameters through additional training on task-specific data, which requires significantly more effort.

With the emergence of long-context LLMs, ICL has shown improved generalization over finetuning [7]. These extended context capabilities also enable integrations such as cache-augmented generation (CAG) and multi-step reasoning with tool use for complex tasks [8]. CAG involves preloading relevant data into the model’s context and caching parameters, allowing inference without further retrieval. This is especially effective when the knowledge base is compact and well-defined. We utilize the concept of preloading by providing the full text of a publication in the first message, which can save processing cost in our multi-turn workflows. As we compare multiple models through the OpenRouter API, the caching is ultimately in the hands of the respective providers.

The strength of modern LLMs as sophisticated, stochastic text generators entails their biggest downside: the “hallucination” of non-existent entities or factually incorrect statements. In a biomedical context, made-up proteins or gene-disease links are hard to spot, even for experts. Hence a mechanism is needed to reconcile predictions with reality, which is known as grounding [9]. Our approach exploits the increased capability of current models to reliably use external “tools”, by verifying LLM-suggested terms with the help of the PubTator 3 API [10, 11]. Because it uses a controlled vocabulary, we also overcome the difficulties of synonyms or other notation-related errors.

A four-step methodology for annotation was established in Giuliani et al 2025 [10], where the combined approach integrating in-context learning, providing said context and using external tools was first investigated. In this first study, predicted annotations achieved an average precision of 98% after verification by domain experts [10]. However, this approach is limited by its implementation with a custom GPT instance using ChatGPT-4o in a browser session [10]. In the present paper, we build upon these insights and assess the feasibility of the four-step workflow for a number of large language models. We implemented a fully automated framework based on API calls, with which we evaluated 14 different LLMs, of which eight models consistently supported reliable execution of the annotation workflow. These eight LLMs were subsequently employed to extract metadata suggestions for six research articles from CRC 1453 “NephGen”. The code for our framework and the data are publicly available.^2^

## Methods

We base the methodology upon our experimental platform for few-shot metadata prediction. It implements a structured approach to information extraction from scientific publications with multi-turn LLM conversations.

The four turns in a single conversation are as follows (Figure 1). First, the model is instructed to extract relevant entities from the full text while disregarding the discussion and bibliography. Second, we direct the model to validate its suggested entities with the provided PubTator 3 tool. Third, we ask the LLM to consider and assign identified entities in context to a predetermined, hierarchical entity structure, using the metadata schema previously developed for CRC “NephGen” [5]. In the final turn, the model is ordered to consolidate the results of all previous steps. An example conversation is available online.^3^ In each turn we expect a JSON list as output, for which we require the model provider to support the Structured Output feature. This guarantees either a JSON output following our specifications or an error message. The sequential output of these lists with generated entity candidates allows the LLM to refine them based on its previous iteration in each step, but also to collate them repeatedly with the publication text. Hence, removing or adding entities can occur in each step while new context is introduced. This enables a “chain-of-thought” refinement towards contextually relevant and grounded entities.

**Figure 1:**
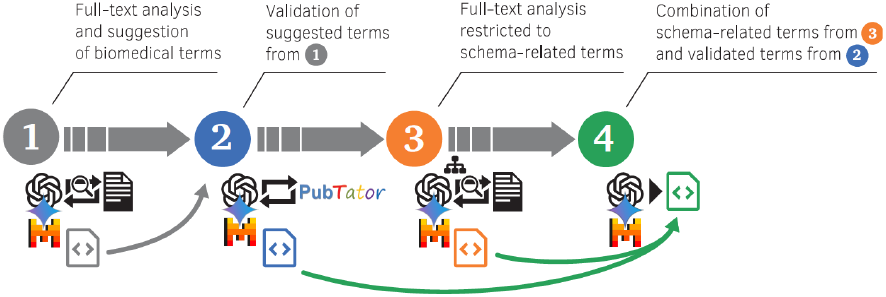
4-step generative workflow.

Turn two requires the Tool Use feature from providers listed on OpenRouter. We define a “PubTator Search” tool, which queries the PubTator 3 API [11], when invoked. Parsed and cleaned results are presented to the LLM. We enforce that the tool has to be used at least once. By storing these calls, we can evaluate if unqueried entity IDs were either hallucinated (not listed in PubTator 3) or inferred from the context with the model’s intrinsic knowledge (listed in PubTator 3).

Subject of our evaluation are six articles from the CRC “NephGen”, published between the first and the second funding period, each authored by a scientist who agreed to participate in a face-to-face interview. This criterion was intentionally chosen, as we believe that the presence of a research data management team member provides the necessary social control to ensure a reliable and diligent evaluation of the automated annotation. During the interviews, all participants thoroughly reviewed the final LLM predictions, classifying them as correct or incorrect, while being allowed to refer to the article’s methods and results sections in case of uncertainty. Each review was limited to five minutes, to balance speed and reliability. Given the impossibility of having a ground truth, we can only calculate the precision of an annotation based on the expert’s judgment.

To estimate precision across articles, we applied meta-analysis methods for single proportions [12]. Given the small number of entities per study, the Freeman-Tukey double arcsine transformation was used to stabilize variances, enhance robustness when proportions approach 1, and accommodate small sample sizes [13]. A random-effects model with the REML estimator was used to account for between-study variance. Differences between LLMs were evaluated using random effects meta regression among transformed values. Differences in the costs, computation time and the total number of biomedical entities between LLMs were estimated with linear mixed models with a random intercept at the study level.

We initially selected 14 LLMs of various parameter counts and modalities (e.g. self-hostable), which met our feature criteria. Note that features such as Tool Use and Structured Output need to be implemented by providers, so availability depends not only on the models capabilities. After preliminary experiments, we removed five models, which were not able to reliably complete full conversations in our workflow. For more details, please refer to supplemental Figure 1. For the interviews, we had to restrict the selection to five models (with six articles each) due to scientists’ time constraints. The three additional models were re-evaluated based on the existing classifications.

## Results

All LLMs performed reasonably well with an overall precision (defined as ratio of correctly identified entities over total number of annotation suggestions) of 91.3% [95%CI 87.4%-94.6%], meaning that the vast majority of predicted biomedical entities were considered correct in the face-to-face interviews. As shown in Figure 2, GPT 4.1, GPT-4o Mini and Gemini 2.0 Flash were associated with the highest precision (95.7%, 97.7% and 93.6%), while GPT 4.1 Nano was associated with the lowest precision (71.9%). None of the differences between LLMs with the highest precision (GPT 4.1, GPT-4o Mini and Gemini 2.0 Flash), however, did reach statistical significance (p=0.516 for the comparison of GPT-4o Mini and Gemini 2.0 Flash, for instance) and can therefore be in the range of randomness.

**Figure 2:**
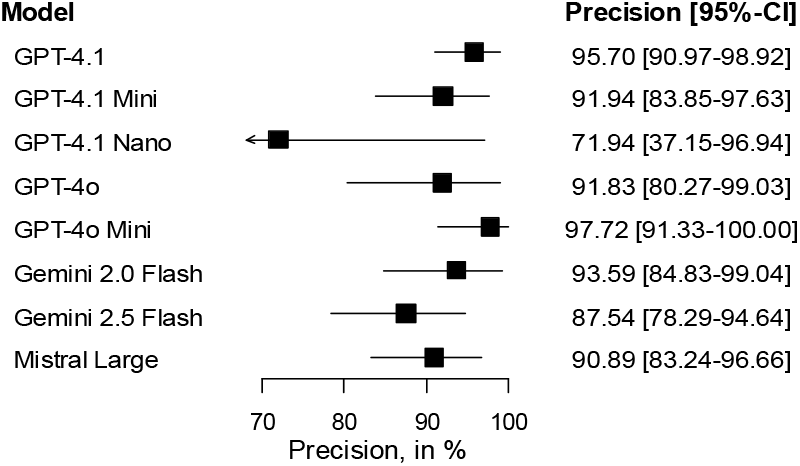
Precision of the different LLMs

Overall, Gemini was the most diligent when it came to proposing biomedical entities. The mean number of correct entity annotations here was 47.5 and 60.8 for Gemini 2.0 Flash and Gemini 2.5 Flash, respectively. GPT-4o and GPT-4o Mini, in contrast, only proposed a mean of 22.8 and 21.0 entities per article (Table 1). As can be seen in Figure 3, the total number of LLM-suggested biomedical entities was not correlated with the precision of the respective approach (p=0.710). On the contrary, both GPT-4.1 and Gemini Flash 2.0 have a high rate of suggested entities with high precision (Table 1).

**Table 1:**
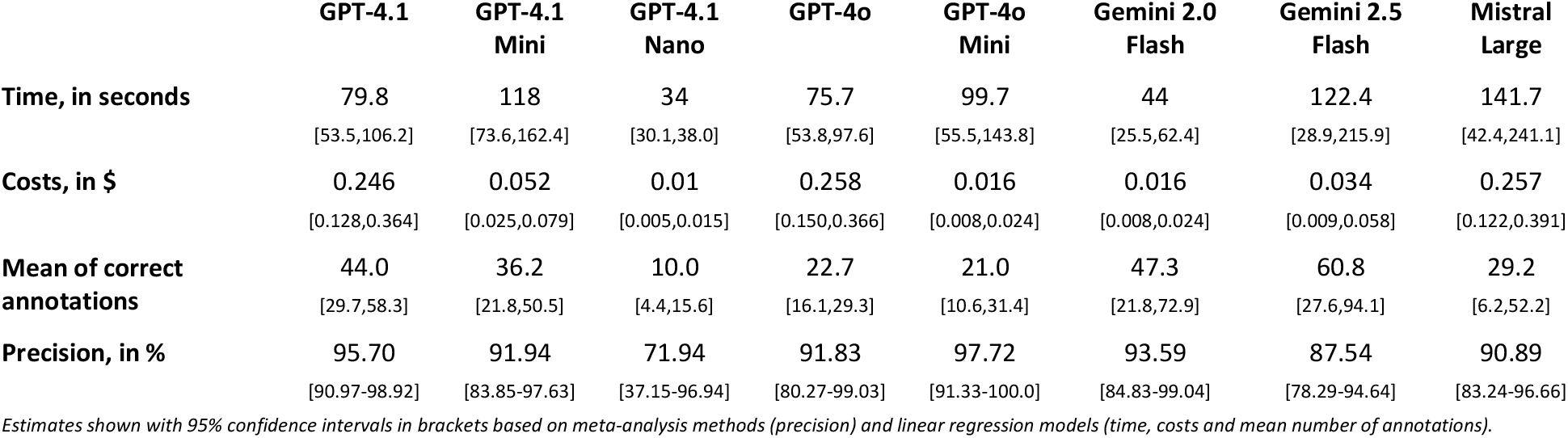
Results of the 4-step approach

**Figure 3:**
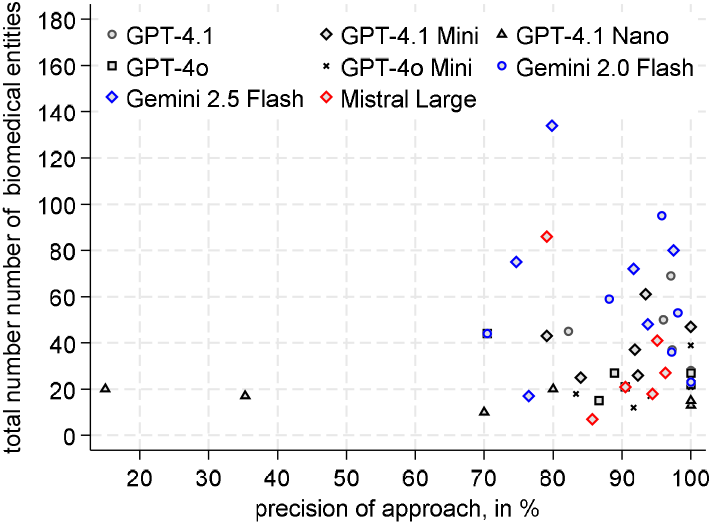
Precision of the different LLMs

In Figure 4, we see the Pareto frontier of our approaches with respect to precision and number of correct predictions. In this case, GPT 4.1, GPT-4o Mini, Gemini 2.0 Flash and Gemini 2.5 Flash are Pareto efficient compared to the other approaches.

**Figure 4:**
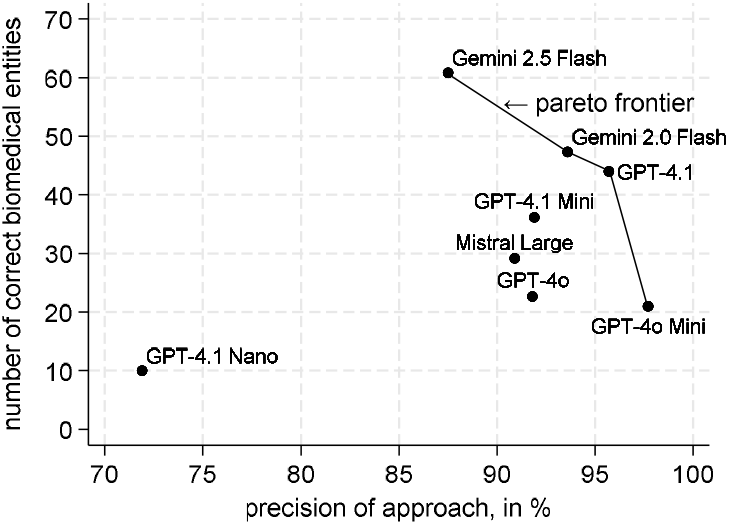
The Pareto frontier with respect to precision and number of correct predictions

Besides the scientific relevance measured by the interview, we evaluated the grounding with PubTator 3 in step 2 of the workflow. Supplemental Figure 1 illustrates the efficacy of tool use for different model groups. As in the interview verification, the Gemini models predict a consistently high number of grounded entities with hallucinations occurring only for Gemini 2.5 Flash. Despite the naming scheme, performance did not increase from 2.0 to 2.5 or from Flash to Pro. Such improvements, however, can be seen for the GPT model class, where 4.1 and its Mini variant outperformed the older 4o models. The distillation to the Mini variant shows even a slightly better entity identification and the list shrunk significantly for GPT-4.1 Nano (see supplemental Figure 2). Overall, the rate of false positives was low for models that could successfully complete a conversation.

There were considerable differences in the costs of the LLM execution (Figure 5). Here, Gemini 2.0 Flash and GPT-4.1 Nano were associated with the lowest costs. At the time of the study, the costs here were $0.1 per million input tokens and $0.4 per million output tokens, respectively. On average, this resulted in costs of only $0.014 and $0.010 for the execution of the 4-step workflow with Gemini 2.0 Flash and GPT-4.1 Nano, respectively. GPT-4o Mini, Gemini 2.0 Flash and GPT-4.1 Mini, were nearly as favorable, with million token costs (input/output) of $0.15/$0.6, $0.15/$0.6 and $0.4/$1.6, respectively. GPT-4.1, GPT-4o and Mistral Large were in the upper range in terms of cost ($2/$8, $2.5/$10 and $2/$6, respectively) and all led to correspondingly higher costs per annotation. As shown in Figure 5, GPT-4o Mini is the sole Pareto efficient model with respect to interview precision and cost. Interestingly, the cost differences between the two models with the highest precision (GPT-4.1 and GPT-4o Mini) was more than 10fold ($0.246 vs. $0.016, p<0.001).

**Figure 5:**
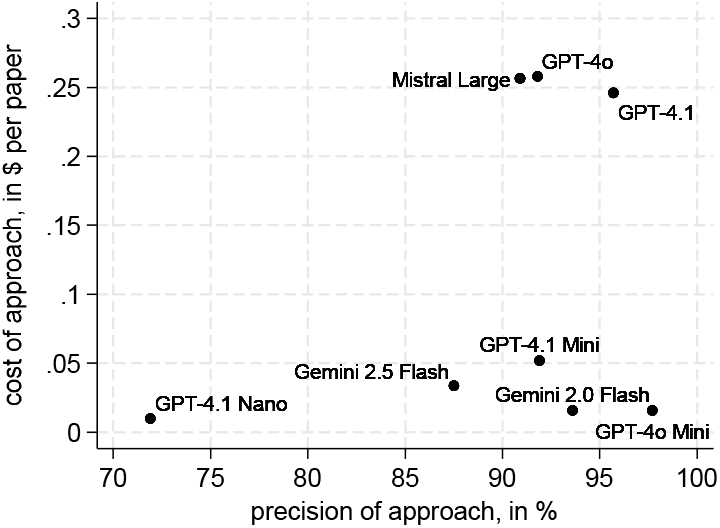
The precision and costs of LLMs

The same is true for the duration of the LLM processing. This was measured as the time between the request for every step and the corresponding response, excluding the tool call communication. GPT-4.1 Nano and Gemini 2.0 Flash were associated with the fastest execution time. As shown in Figure 6, Mistral Large needed, on average, more than three times as long as Gemini 2.0 Flash. The Pareto frontier of our approaches with respect to precision and duration shows that GPT-4.1 Nano, Gemini 2.0 Flash, GPT-4.1 and GPT-4o Mini are Pareto efficient compared to the other approaches.

**Figure 6:**
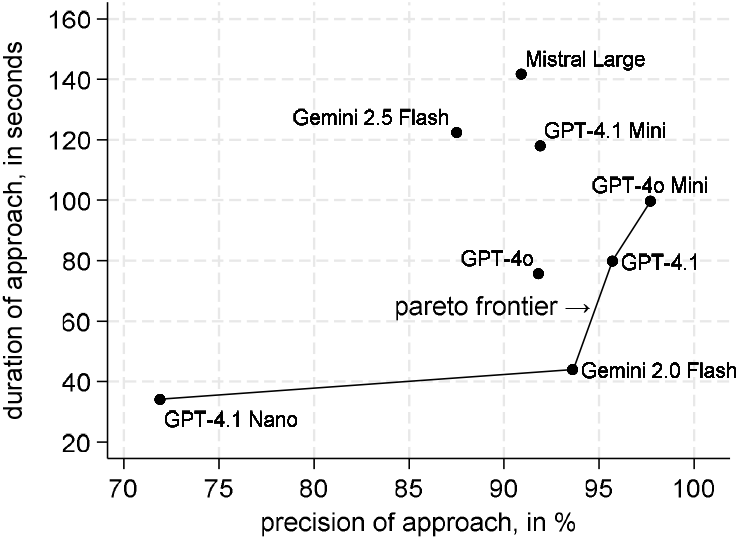
The Pareto frontier with respect to precision and duration of the LLM processing

## Discussion

Overall, the investigated LLMs predicted biomedical entities quite reliably. Only GPT-4.1 Nano showed some downward outliers in its precision. We had previously observed similar results with Mistral Small and Mistral Medium, which, when run successfully, appeared less reliable than Mistral Large. In addition, both Mistral Small and Mistral Medium regularly failed to complete conversations, which is why they were not included in the interview evaluation. GPT-4.1 Nano performed the workflow reliably, but the results of at least one run suggest that the model is too small for the task at hand. Surprisingly, we found that the larger GPT 4.1 and GPT-4o ran faster than their “mini” counterparts, despite having a larger number of identified biomedical entities. In our opinion, this can only be explained by resource prioritization for the significantly more expensive models by their respective providers. An exact evaluation of duration and reliability is difficult, because the providers can change the experimental setting without notifying the users. At several points our experiments had to be delayed, because severe bugs regarding tool use and structured output needed to be fixed in the providers’ SDKs.

The multidimensional decision problem of LLM choice was taken into account by means of the Pareto frontiers. The precision was chosen as the primary decision criterion. The total number of correctly predicted biomedical entities can be seen as a secondary criterion. The large gap between GPT-4.1, Gemini Flash 2.0 and GPT-4o Mini shown here does not make the final decision easy. The speed and costs of the respective approach were considered tertiary in this study. Collaborative research centers focus on quality and processing time is of marginal relevance. In a public implementation of the presented approach, energy consumption [14– 16] and cost-effectiveness of the respective LLMs could become more important [17, 18]. Here, self-hosting locally available open-source LLMs would be more cost-effective than the LLM-as-a-Service strategy we have chosen [19]. Unfortunately, none of the self-hostable LLMs reached a sufficient precision.

We may conclude that the applied LLM annotation works well and has the potential to accelerate the annotation of papers and datasets. A key challenge, however, is distinguishing between articles and their underlying datasets. While metadata prediction is article-based, annotations should target datasets to facilitate their reuse. Beside possibly conflated information about the respective datasets, the publication also addresses the scientific context it contributed to, which entails the use of entities that are irrelevant to data annotation. To mitigate this discrepancy, we instructed the LLMs to disregard the discussion and bibliography sections during the prediction phase (step 1). Despite this measure, some incorrect predictions suggest that this part of the approach may not have been fully effective. Nonetheless, we consider it essential to apply and refine entities using the full text of each article, rather than limiting the input to selected sections. This strategy appears to be a more viable path towards establishing a fully automated annotation process, because it provides relevant information to the LLM instead of censoring the input.

As we have shown, the number of suggested biomedical entities varied substantially across LLMs, which makes the choice of a specific LLM elementary. For now, we highly recommend that LLM-assisted metadata annotations should undergo human review before publication (“human-in-the-loop”). This could be both technically enforced and systematically integrated into the workflow.

A further limitation of the approach is the inability to identify biomedical entities that were not predicted by the LLMs. In other studies on natural language processing with LLMs, false negatives are commonly taken into account when calculating performance metrics such as accuracy, recall, and the F1 score. However, in the present context, where the primary objective is to maximize the number of correct annotations while minimizing the proportion of false positives, this approach is not feasible [10]. The key limitation lies in the fact that the number of potential annotations per dataset does not have a clearly defined upper bound. While it would, in principle, be possible to ask authors during face-to-face interviews which biomedical entities they believe are missing, obtaining this information would require considerable additional effort. Nevertheless, since our approach deliberately guides the LLM by providing it with the predefined CRC metadata schema, it at least ensures that the generated annotations are not only contextually appropriate for the given paper but, more importantly, are also relevant in relation to the CRC framework.

## Data Availability

All data produced in the present work are contained in the manuscript

---

**Supplemental Figure 1:**
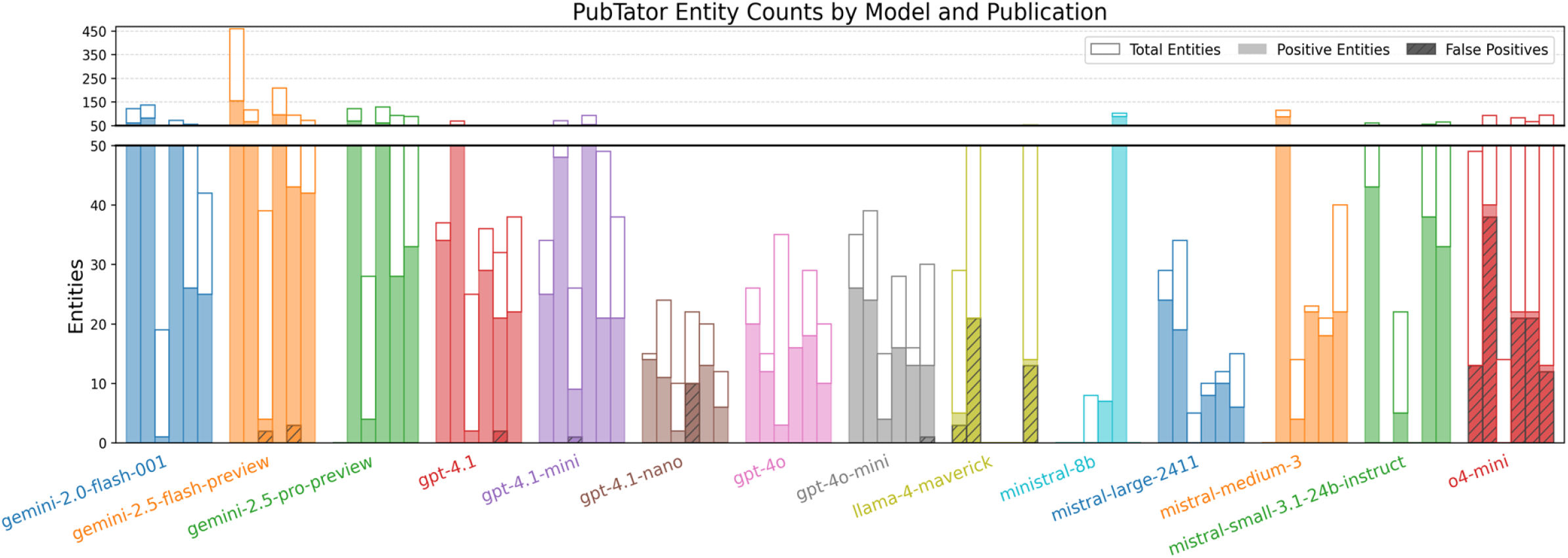
The cardinality of the entity set after step 2 for each model and each publication. Each bar indicates how many entities were listed in total (step 1), positively predicted to be in PubTator 3 with an ID (step 2) and how many were hallucinations and do not exist in the database. Missing bars indicate failed. Finally, we were able to build a stable pipeline for operationalizing the described four-step approach with eight LLMs: Google Gemini 2.0 Flash 001, Gemini 2.5 Flash (2025-04-17), OpenAI GPT 4o (2025-03-26), OpenAI GPT 4o mini (2024-07-18), Mistral Large 2411, OpenAI GPT 4.1 mini (2025-04-14), OpenAI GPT 4.1 nano (2025-04-14) and OpenAI GPT 4.1 (2025-04-14).

**Supplemental Figure 2:**
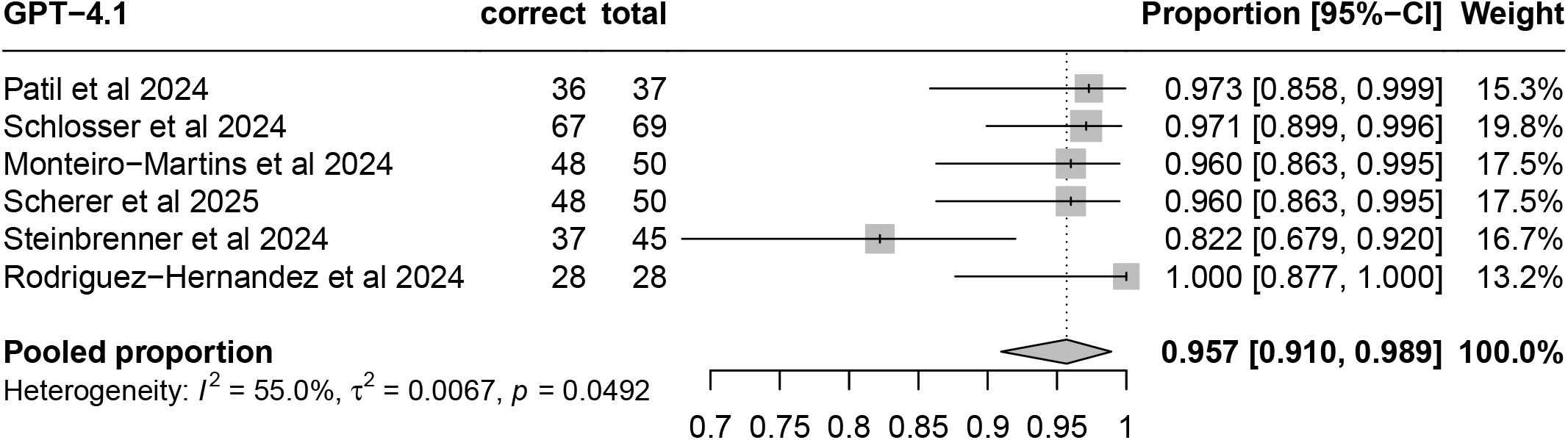
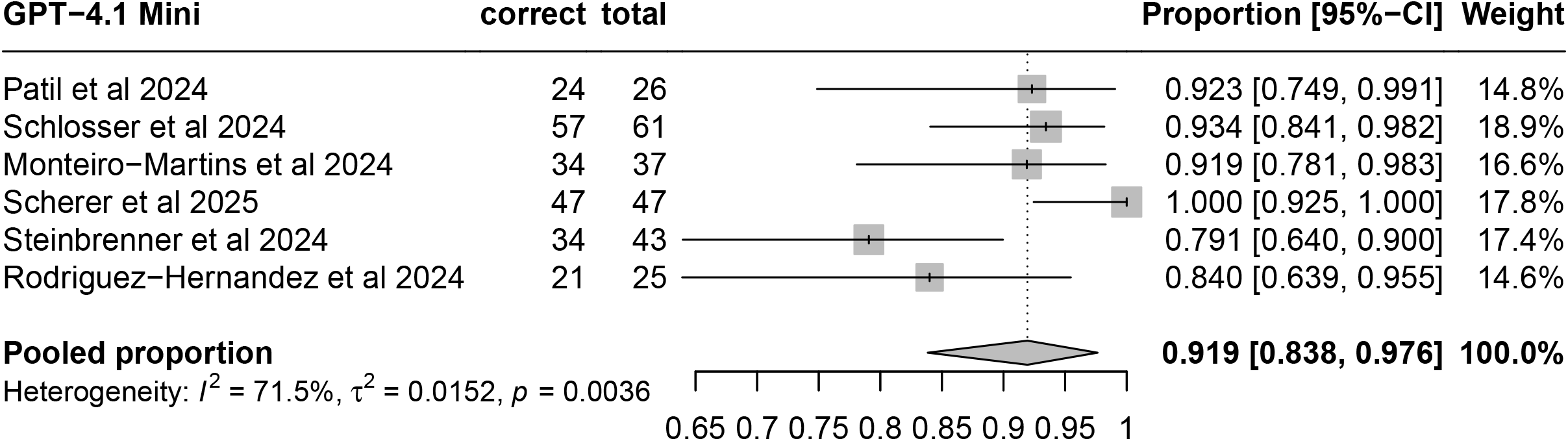
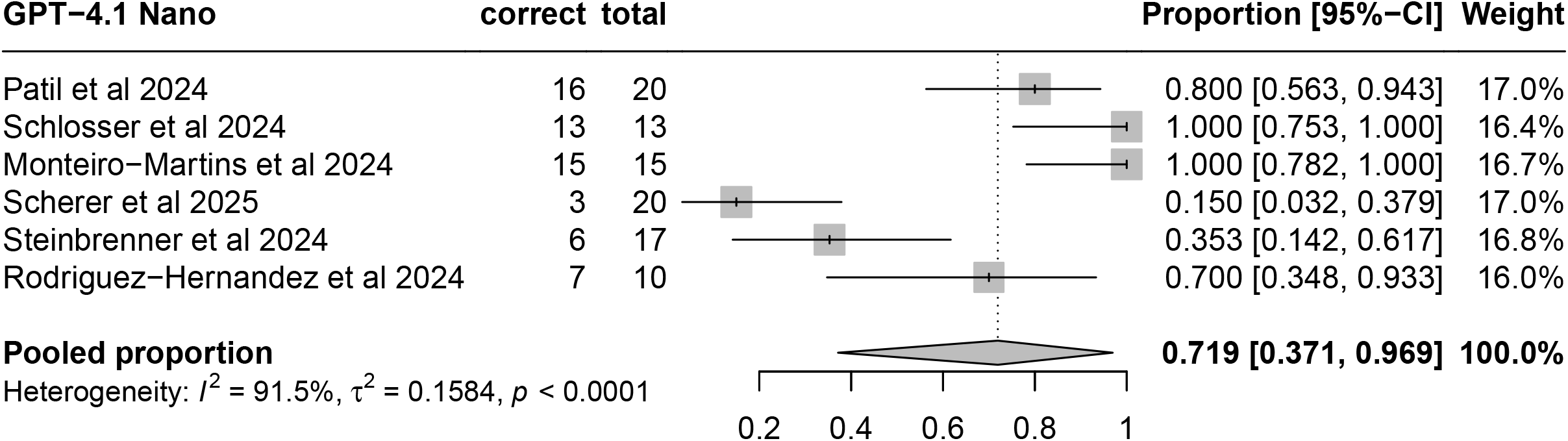
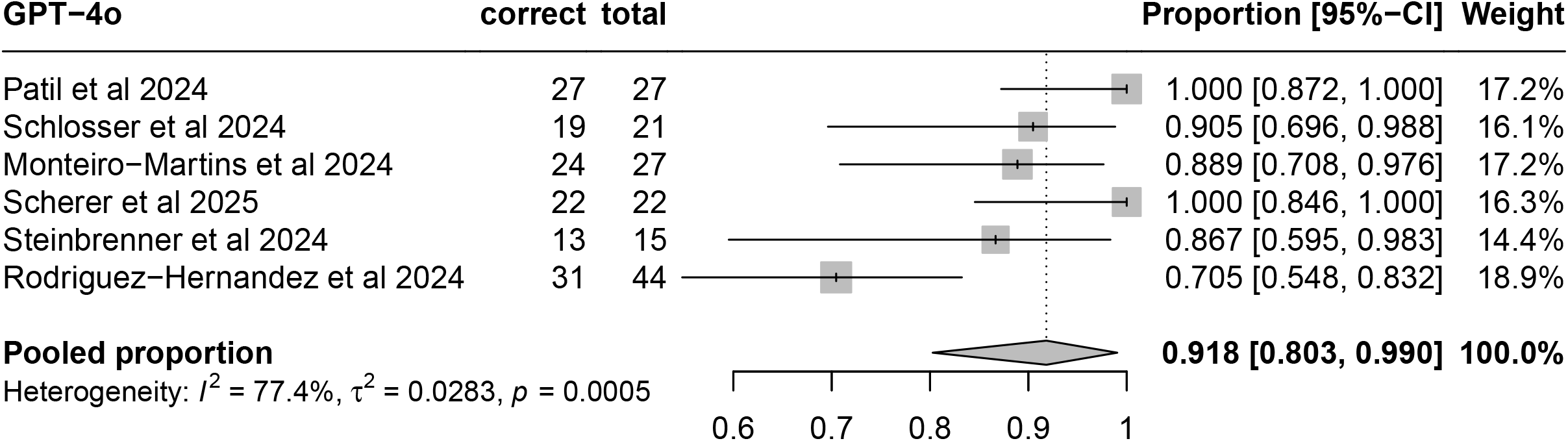
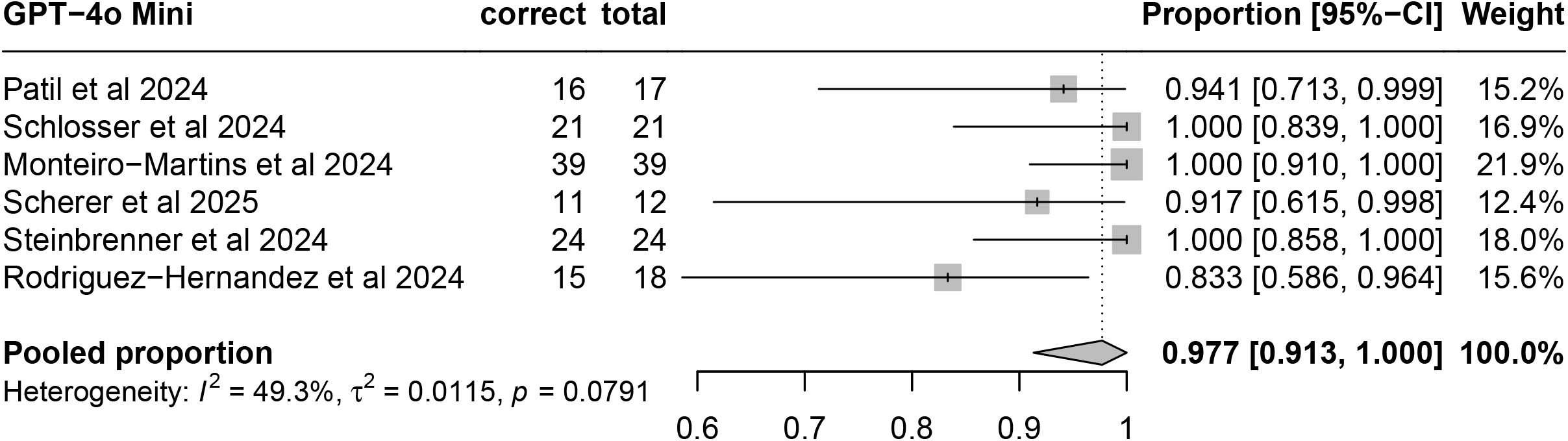
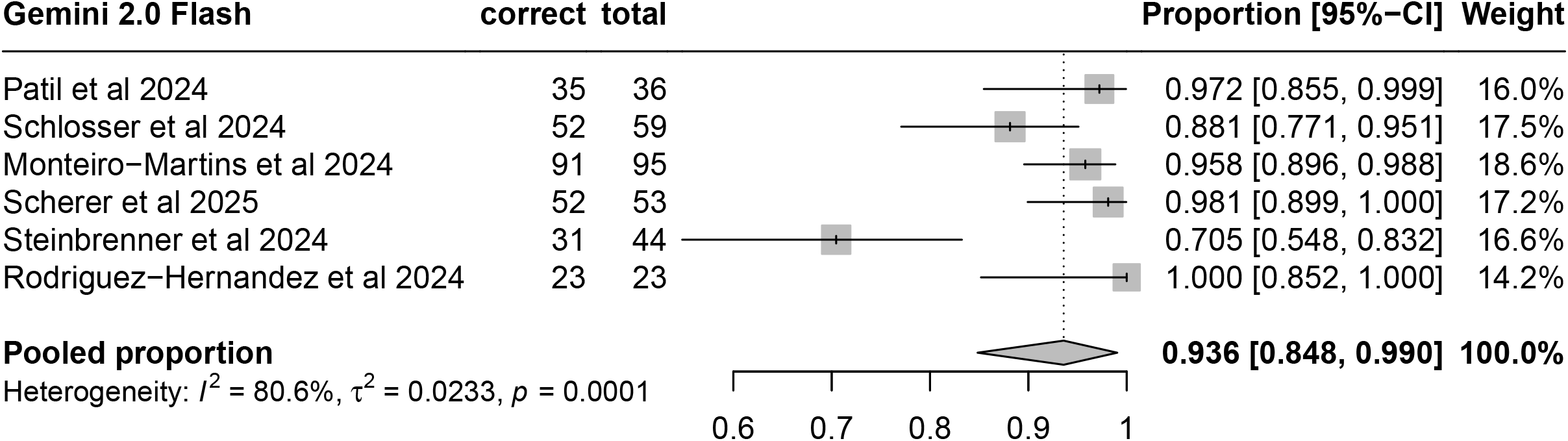
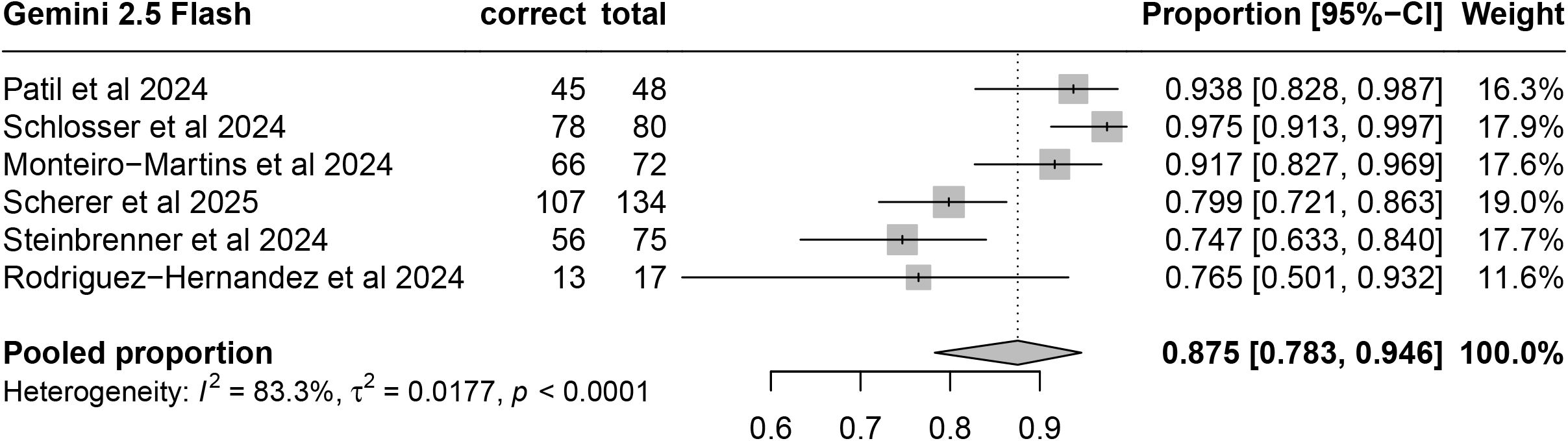
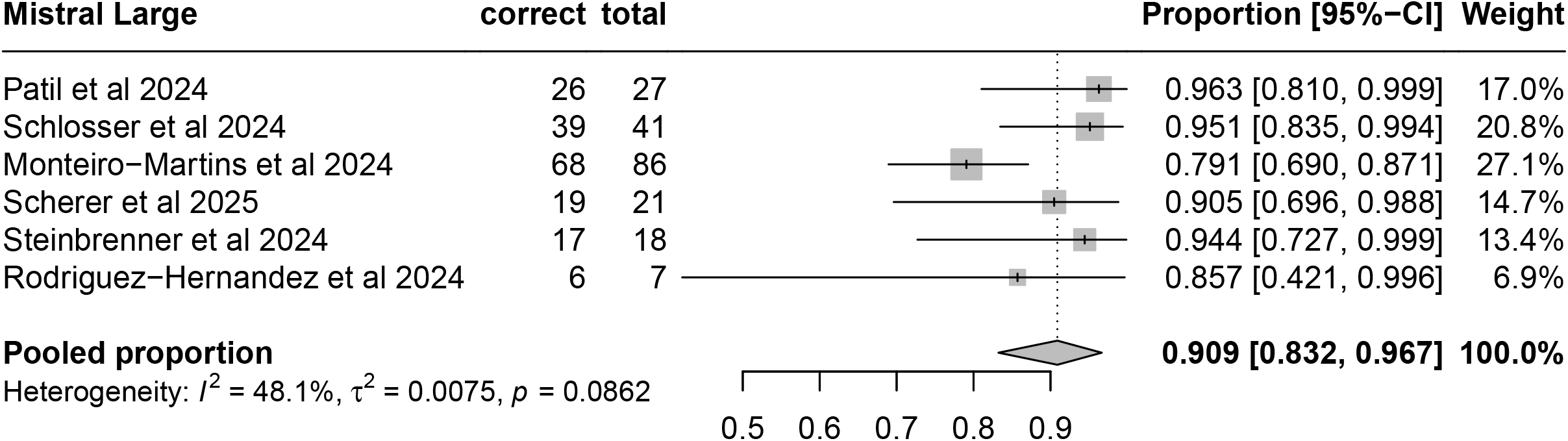
Results of the meta-analyses

## Footnotes

2 https://github.com/watterm/llm-metadata-annotation

3 https://watterm.github.io/llm-metadata-annotation/example.html

